# TwinsMX: Exploring the Genetic and Environmental Influences on Health Traits in the Mexican Population

**DOI:** 10.1101/2023.09.20.23295527

**Authors:** B. García-Vilchis, T.V. Román-López, D. Ramírez-González, X. López-Camaño, V. Murillo-Lechuga, X. Díaz, I.C. Sánchez-Moncada, I.M. Espinosa-Méndez, D. Zenteno-Morales, Z.X. Espinosa-Valdes, S. Pradel-Jiménez, A. Tapia-Atilano, A.V. Zanabria-Pérez, F. Livas-Gangas, O. Aldana-Assad, U. Caballero-Sánchez, C.A. Dominguez-Frausto, M.E. Rentería, A. Medina-Rivera, S. Alcauter, A.E. Ruiz-Contreras

**Author notes:** Corresponding authors: **Sarael Alcauter, Ph.D.**, Instituto de Neurobiología, Universidad Nacional Autónoma de México, **Alejandra E. Ruiz-Contreras, Ph.D.**, Facultad de Psicología, Universidad Nacional Autónoma de México. These authors jointly led and supervised the project (M.E. Rentería, A. Medina-Rivera, S. Alcauter, A.E. Ruiz-Contreras). These authors contributed equally to this work (B. García-Vilchis, T.V. Román-López).

## Abstract

TwinsMX registry is a national research initiative in Mexico that aims to understand the complex interplay between genetics and environment in shaping physical and mental health traits among the country’s population. With a multidisciplinary approach, TwinsMX aims to advance our knowledge of the genetic and environmental mechanisms underlying ethnic variations in complex traits and diseases, including behavioral, psychometric, anthropometric, metabolic, cardiovascular, and mental disorders. With information gathered from over 2800 twins, this article updates the prevalence of several complex traits; and describes the advances and novel ideas we have implemented such as magnetic resonance imaging. The future expansion of the TwinsMX registry will enhance our comprehension of the intricate interplay between genetics and environment in shaping health and disease in the Mexican population. Overall, this report describes the progress in the building of a solid database which shall allow to study complex traits in the Mexican population, valuable not only for our consortium but for the worldwide scientific community by providing new insights of understudied genetically admixed populations.

## Introduction

Twin studies allow the study of the genetic and environmental influences on human complex traits and diseases. Researchers can gain insights into the relative contributions of genetics and environment by comparing the similarities and differences in specific traits between monozygotic (who are ∼100% similar in their genomes) and dizygotic twins (who are similar in around 50% on their genomes); both sharing environmental factors, particularly during childhood (Hur & Craig, 2013). By comparing similarities and differences between these twin types, researchers can unravel the interplay between genetic and environmental factors in determining specific traits and diseases (Polderman et al., 2015).

Over the years, twin studies have helped to elucidate the underlying causes of many phenotypes and diseases and have provided valuable insights for the development of new therapies and treatments (Petrill et al., 2013; Plomin et al., 2014; Luciano et al., 2015; Verweij et al., 2018; de Moon et al., 2019). Moreover, they have played a pivotal role in shaping prevention strategies, which have the potential to enhance public health practice (Rogowski et al., 2009) benefiting the Mexican population.

Numerous nations, mostly in Europe, North America, and Asia, have established twin registries (Chen & Liang, 2013; Derom et al., 2016; Yoon-Mi et al., 2019). Yet, relatively few genetic studies have been conducted in admixed populations. This is a significant gap in our knowledge because these communities possess a unique genetic architecture due to their socio-demographic history (Petrill et al., 2013). Without comprehensive data from these populations, we are potentially overlooking unique genetic contributions to health and disease, and it may limit the precision of medical interventions based on genetics. Therefore, by studying these populations, we might uncover new genetic variations that influence how individuals respond to illnesses and medications (Chen & Liang, 1999; Dupuis et al., 2004). Moreover, including underrepresented populations in genetic studies can foster social justice and equity in access to precision medicine, as these communities have often been excluded from genetic research, potentially limiting their ability to benefit from advancements in medical science and clinical applications (Martin et al., 2019; Peterson et al., 2019; Wojcik et al., 2019). The Mexican population, a product of genetic admixture, displays varying degrees of genetic ancestry composition depending on the region. Generally, it is estimated to range from 56% to 90% for indigenous, from 10% to 38% for European ancestry, and around 5% and 2% for African and Asian ancestry, respectively (Juárez-Cedillo et al., 2008; Martinez-Fierro et al., 2009; Ruiz-Linares et al., 2014; Bodner et al., 2021; De Oliveira et al., 2023).

In addition to genetic variation between populations, Mexico has a higher prevalence of particular diseases. For instance, metabolic-related disorders like obesity, diabetes, and hypertension are more common when compared to populations with European ancestry. In Mexico, 14.3% of adults suffer from diabetes, compared to 8.8% in Spain, 7.1% in Italy, and 6.9% in the United Kingdom (WHO, 2019). In comparison with other populations, cardiovascular diseases cause about 30% of all deaths in Mexico (Secretaría de Salud de México, 2016). This figure compares to 27% in France (Tuppin et al., 2016), and 34.8% in Italy (Cortesi et al., 2021). On the other side of the world, China experiences an even higher rate, with cardiovascular diseases accounting for 40% of all deaths (Liu et al., 2019), whereas Japan has 28% of deaths from the same cause (Ikeda et al., 2009). On the other hand, considering mental health in Mexico, the prevalence of depression is 18.3% and anxiety is 14.3% in adults (Medina-Mora et al., 2018; Agudelo-Botero et al., 2021), whereas, it is 3.3% and 4.3%, respectively, in Europe (Santomauro et al., 2021). These data highlight that studies developed in populations with European or Asian ancestries do not necessarily fit public health issues in the Mexican population.

The aims of this article are, firstly, presenting an update on the TwinsMX registry, which was established in 2018 (Leon-Apodaca et al., 2019). This update details the prevalence of various complex traits observed in monozygotic and dizygotic twins. Secondly, to inform about recent developments such as the inclusion of brain magnetic resonance imaging (MRI) scans for twins. Thirdly, shedding light on the challenges and limitations encountered in the TwinsMX registry and to describe strategies for future growth. Finally, the article aims to stimulate a deep understanding on the complexities involved in the study of genetic traits in the Mexican population.

## Materials and methods

### Experimental Design and Progress to Date

#### Participants

Mexican twins, triplets, and other multiple-birth participants have been recruited by means of demographic campaigns launched across the country. Twins of different ages were contacted by means of social media (Facebook, Twitter, and Instagram), and newspapers, as well as nursing homes, large hospitals, and educational institutions. Adult twins over 18 years old and underaged children, with the assistance and authorization of their primary caregiver, completed an online survey at www.twinsmxofficial.unam.mx. Subjects provided their written consent to participate in the registry. The procedures executed in TwinsMX have been revised and approved by the Research Ethics Committee of the Institute of Neurobiology at UNAM.

#### Data collection

Several measures are being collected, like anthropometric(i.e. height and weight) and epidemiological information (i.e., medical history, lifestyle, quality of life, mood, cognitive function, and nutritional status). Some of these surveys have been previously carried out on the Mexican population and currently serve as control and updating for the project (Ruiz-Contreras et al., 2012; Alcauter et al., 2017; Gracia-Tabuenca et al., 2018). Participants are requested to consent for future contact, to participate in later stages of the study, and to express their will to donate a DNA sample for genotyping. Moreover, twins will be invited to participate in a magnetic resonance imaging (MRI) session that includes a cognitive evaluation (*e*.*g*., working memory, planning, and deductive reasoning) with the Creyos platform (https://creyos.com/; Gleichgerrcht et al., 2019), and the acquisition of anatomical and functional magnetic resonance imaging (fMRI), and magnetic resonance spectroscopy of the medial frontal and parietal lobes to estimate the concentration of metabolites, including gamma aminobutyric acid (GABA), glutathione (GSH), glutamate/glutamine (Glx), N-acetylaspartate (NAA), total Creatine (tCr), Choline (Cho) and myo-Inositol (mI). Finally, diffusion weighted imaging is also collected to estimate the white matter integrity and the modeling of the main white matter tracts considering crossing fibers (Grier et al., 2020), using constrained spherical deconvolution or other multifiber methods. This information is securely stored and anonymized in a database on the TwinsMX platform.

The development and maintenance of TwinsMX as Mexico’s National Twin Registry are fully supported by the infrastructure and institutional framework of the *Universidad Nacional Autónoma de México* (UNAM, that may be translated as the National Autonomous University of Mexico). The Institute of Neurobiology, the International Laboratory for Human Genome Research, both in the city of Querétaro, and the School of Psychology in Mexico City are where the main research team is headquartered. Similar resources at UNAM, such as the National Laboratory of Magnetic Resonance Imaging and the National Laboratory of Advanced Scientific Visualization, also offer TwinsMX infrastructure and assistance.

#### Data Collection Platform

Participants answer a series of online surveys and self applied psychological inventories using the Research Electronic Data Capture (REDCap) platform. REDCap facilitates the integration of surveys and document collections in research projects while streamlining data administration for statistical analysis (Busch et al., 2015; Wright, 2016; Evers & Schouten, 2018; Harvey, 2018). The “Unique Population Registry Code” (CURP by its initials in Spanish), a valid Mexican identity number, must be entered by each TwinsMX participant, this ID is used as their identification number to enter the platform and due to its composition that includes information on the birth date and place of birth, it can be used to match each twin to their siblings.

#### Zygosity assessment.

In the TwinsMX registry, twin zygosity has been determined by twins’ self-report. When the two twins disagreed, gave contradictory answers, or neither twin provided a response, zygosity was labeled as unknown (UZ). If only one twin responded, only this answer was used to determine the zygosity (Ordoñana et al., 2013). Thus, zygosity was classified as monozygotic males (MZM), monozygotic females (MZF), dizygotic males (DZM), dizygotic females (DZF), the dizygotic opposite sex (DZO), and unknown zygosity (UZ).

## Results

### Recruitment to date

Currently, in the TwinsMX registry there are over 2,826 twins from all states of the country. The proportion of registrations by state is represented in **Figure 1**. Mexico City and the State of Mexico, the two states with the most significant number of habitants in the country accounting for over 21 million of persons, are the sites with more registries.

**Figure 1.**
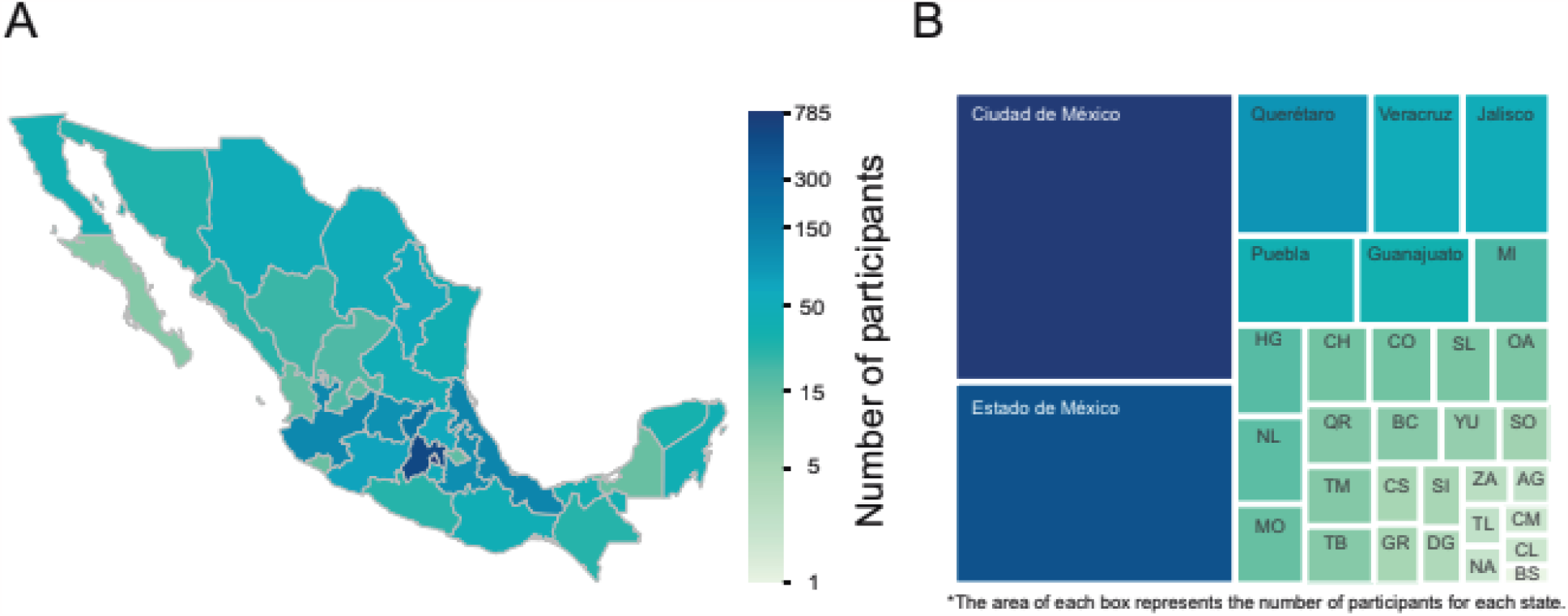
The geographic distribution of the registered twins. (A) The map shows the progress of registrations in each state of the Mexican Republic, the number of registrations is coded with a color scale shown in the central bar. Because of the imbalance in the twins’ participation in geographical terms, the color scale is on a logarithmic scale. (B) The graph represents the record advance in terms of the area of each rectangle, and the color code is maintained. For reference, the codes shown in the figure are: AG: Aguascalientes, BC: Baja California, BS: Baja California Sur, CM: Campeche, CS: Chiapas, CH: Chihuahua, CO: Coahuila, CL: Colima, DG: Durango, GR: Guerrero, HG: Hidalgo, MI: Michoacán, MO: Morelos, NA: Nayarit, NL: Nuevo León, OA: Oaxaca, QR: Quintana Roo, SL: San Luis Potosí, SI: Sinaloa, SO: Sonora, TB: Tabasco, TM: Tamaulipas, TL: Tlaxcala, YU: Yucatán, ZA: Zacatecas (International Organization for Standardization, 2020).

**Figure 2** shows the basic demographic characteristics of twins older than 18 years. **Table 1** provides a more detailed demographic characteristics of twins as a function of age. In the current report, only adults are considered.

**Table 1.** Basic characteristics of the twins registered in TwinsMX.

| Information at enrollment | Younger than 18 years<br>n = 204<br>n (%) | 18 years and older<br>n = 2622<br>n (%) |
| --- | --- | --- |
| **Age at enrollment, years** |  |  |
| Range | 0-17 | 18-100 |
| Mean ± SD | 8.7 ± 4.8 | 32.2 ± 9.9 |
| **Gender** |  |  |
| Male | 115 (56.4%) | 684 (26.1%) |
| Female | 89 (43.6%) | 1938 (73.9%) |
| **Region of residence** |  |  |
| Northern Mexico | 45 (22.1%) | 396 (15.1%) |
| Central Mexico | 144 (70.6%) | 2041 (77.8%) |
| Southern Mexico | 15 (7.3%) | 185 (7.1%) |
| **Educational attainment** |  |  |
| Lower than high school diploma | 183 (89.7%) | 42 (1.6%) |
| High school diploma or technical degree | 21 (10.3%) | 382 (14.6%) |
| Degree or higher | 0 (0.0%) | 2198 (83.8%) |
| **Occupation** |  |  |
| Student | 189 (92.6%) | 709 (27.0%) |
| Employed | 2 (1.0%) | 1663 (63.5%) |
| Unemployed | 13 (6.4%) | 220 (8.4%) |
| Retired | 0 (0.0%) | 30 (1.1%) |
| **Family monthly income** |  |  |
| Not specified | 59 (28.9%) | 846 (32.3%) |
| < 450 USD | 49 (24.0%) | 405 (15.4%) |
| 450 < 900 USD | 59 (28.9%) | 802 (30.6%) |
| 900 < 1500 USD | 16 (7.8%) | 304 (11.6%) |
| > 1500 USD | 21 (10.3%) | 265 (10.1%) |

**Figure 2.**
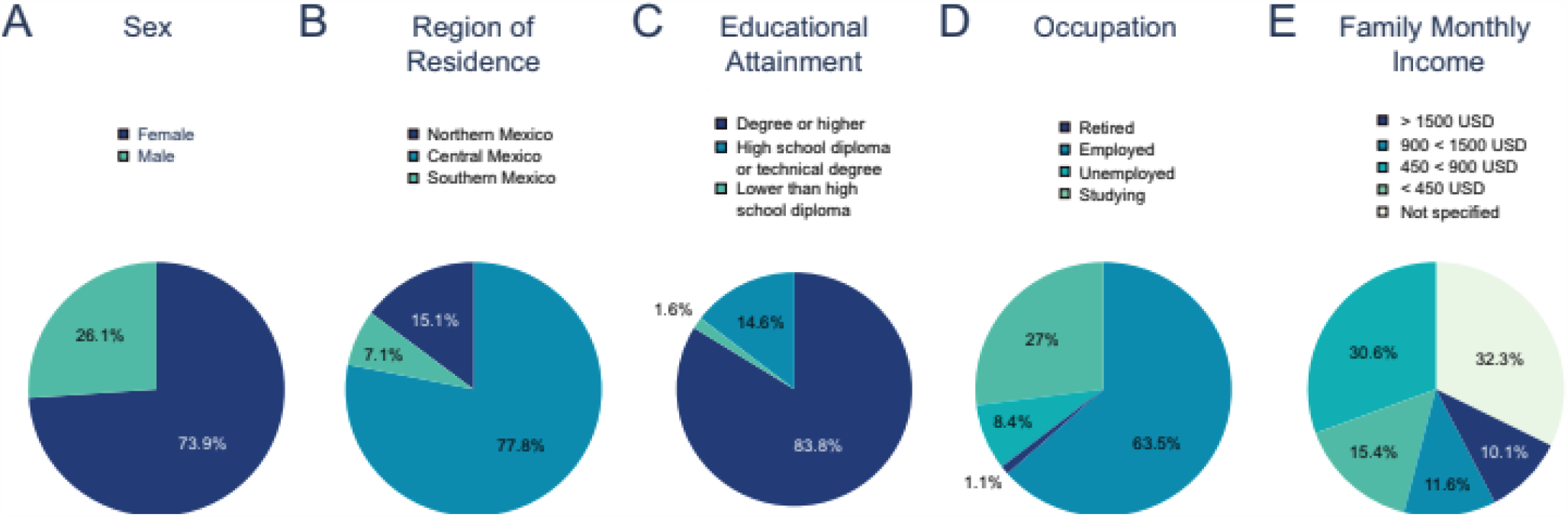
Pie charts illustrating the key demographics of registered twins on TwinsMX. (A) Divided by gender. (B) Percentage of records by geographic region. (C) Proportion of the educational level of the participants. (D) Employment status. (E) Family monthly income. A more detailed version of the data can be found in Table 1.

The study underlines that a significant 73.9% of the records pertain to women, and 77.8% of the participants originate from the nation’s central region (**Figure 1**). A notable majority (83.8%) report holding a bachelor’s degree or higher. Additionally, 63.5% of registered adults are employed, and a minuscule proportion (1.1%) are retired. Around 46% of registries indicate a monthly income below 900 USD (equivalent to less than 10,800 USD per year), whereas 32.3% of participants decided not to disclose their family’s monthly income.

**Figure 3** displays the number of records as a function of age at enrollment, as well as the percentage of records by the self-report zygosity. **Table 2** specifies the number and percentage of twins depending on the type of zygosity, as a function of age.

**Table 2.** Zygosity of registered twins depicted by age group.

|  | MZM | MZF | DZM | DZF | DZO | Unknown zygosity | Total |
| --- | --- | --- | --- | --- | --- | --- | --- |
| Age group | n (row% / col%) | n (row% / col%) | n (row% / col%) | n (row% / col%) | n (row% / col%) | n (row% / col%) | n |
| Under 10 | 25 (22.3% / 5.9%) | 16 (14.3% / 1.6%) | 18 (16.1% / 11.0%) | 17 (15.2% / 3.3%) | 29 (25.9% / 6.3%) | 7 (6.2% / 2.8%) | 112 |
| 10-19 | 24 (14.7% / 5.6%) | 40 (24.5% / 3.9%) | 25 (15.3% / 15.3%) | 32 (19.6% / 6.2%) | 30 (18.4% / 6.5%) | 12 (7.4% / 4.9%) | 163 |
| 20-29 | 141 (11.8% / 33.2%) | 432 (36.3% / 42.4%) | 63 (5.3% / 38.2%) | 229 (19.2% / 44.6%) | 217 (18.2% / 46.9%) | 109 (9.1% / 44.3%) | 1191 |
| 30-49 | 146 (17.1% / 34.6%) | 325 (38.1% / 31.9%) | 31 (3.6% / 18.8%) | 151 (17.7% / 29.4%) | 118 (13.8% / 25.5%) | 82 (9.6% / 33.3%) | 853 |
| 40-49 | 61 (18.1% / 14.4%) | 138 (40.9% / 13.5%) | 17 (5.0% / 10.3%) | 53 (15.7% / 10.3%) | 52 (15.4% / 11.2%) | 16 (4.7% / 6.5%) | 337 |
| 50-59 | 15 (11.5% / 3.5%) | 48 (36.9% / 4.7%) | 8 (6.2% / 4.9%) | 27 (20.8% / 5.3%) | 14 (10.8% / 3.0%) | 18 (13.8% / 7.3%) | 130 |
| 60-69 | 8 (24.2% / 1.9%) | 18 (54.5% / 1.8%) | 0 (0.0% / 0.0%) | 3 (9.1% / 0.6%) | 2 (6.1% / 0.4%) | 2 (6.1% / 0.8%) | 33 |
| Over 70 | 5 (71.4% / 1.2%) | 1 (14.3% / 0.1%) | 1 (14.3% / 0.6%) | 0 (0.0% / 0.0%) | 0 (0.0% / 0.0%) | 0 (0.0% / 0.0%) | 7 |
| Total | 425 | 1018 | 163 | 512 | 462 | 246 | 2826 |

**Figure 3.**
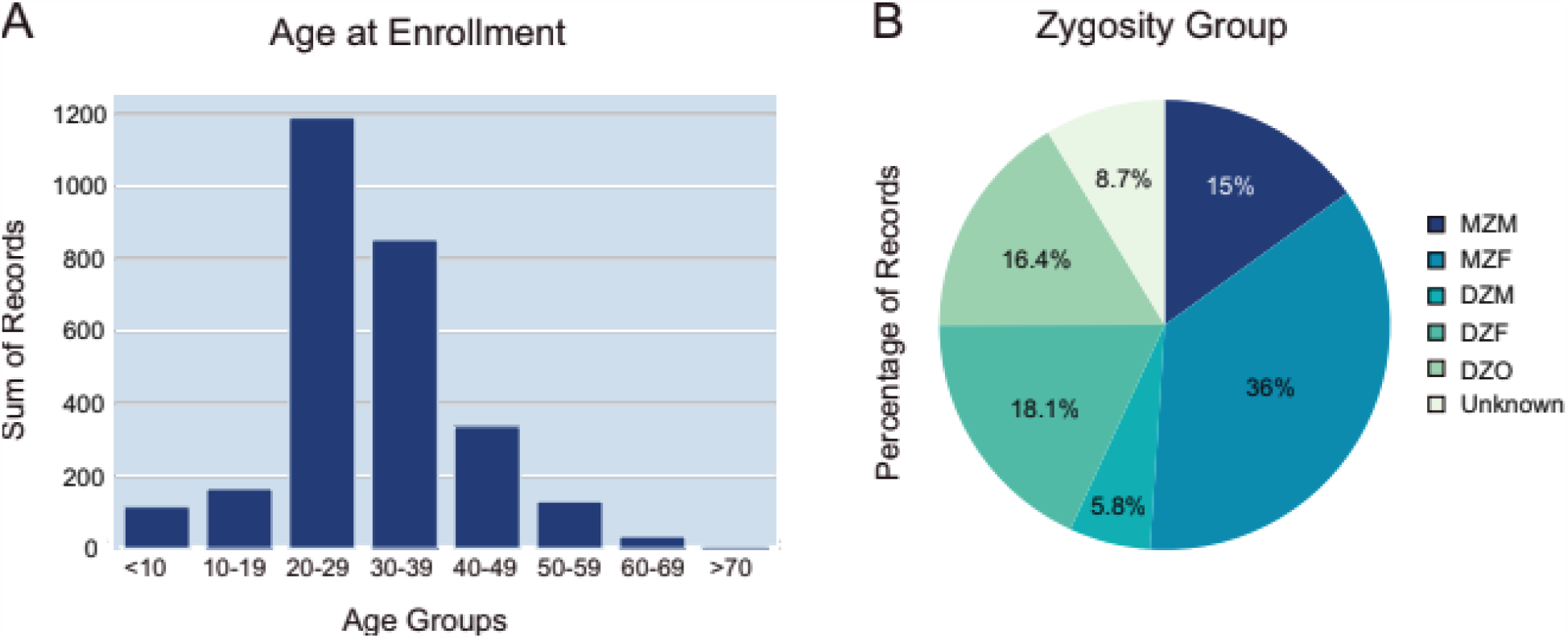
Age and zygosity distribution among the participants. Adults between the ages of 20 and 40 make up a major portion of the histogram in (A). (B) The share of records that are categorized by zygosity. Monozygotic twins and female participants are more common.

Given that there are around 9% of records whose zygosity is awaiting validation, **Table 2** shows that monozygotic twins (n=1443 individuals, MZ) exceed dizygotic twins (n=1137 individuals, DZ). The age range with the greatest number of records is 20-29 years (42.1%), followed by the 30-39 age range (30.2%), for each zygosity group. This age pattern is repeated in all zygosity categories.

According to **Figure 4**, the number of registries has been rising over time. To do follow-up research and even future longitudinal studies, it is expected to actively encourage parents of twins to register their children and participants to update their information.

**Figure 4.**
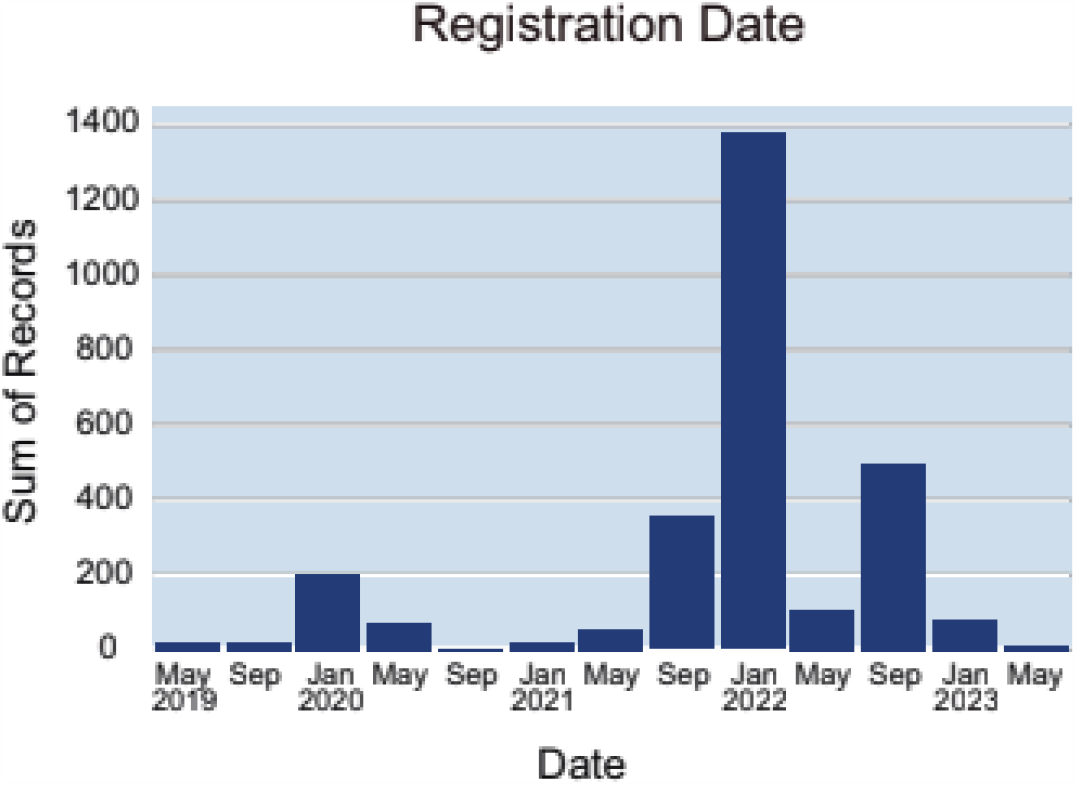
Number of Registered Participants per Quarter. Campaigns have been carried out in the main states of the country to recruit twins. Visits to national television networks usually provide a large number of records in the following weeks.

### Preliminary survey results

The ten physical and mental conditions with the highest frequency among the twins taking part in the study are listed in **Tables 3 and Table 4** as a function of zygosity group and gender.

**Table 3.** The physical conditions with the greatest occurrence rates among twins recorded on TwinsMX.

|  | Myopia | Astigmatism | Gastritis | Colitis | Overweight | Obesity | Acne | Allergic Rhinitis | Gastroesophageal reflux | Haemorrhoids |
| --- | --- | --- | --- | --- | --- | --- | --- | --- | --- | --- |
| Zygosity group | n (%) | n (%) | n (%) | n (%) | n (%) | n (%) | n (%) | n (%) | n (%) | n (%) |
| MZM | 173 (12.2%) | 141 (12.1%) | 106 (12.2%) | 69 (9.0%) | 83 (12.5%) | 37 (13.6%) | 75 (12.3%) | 42 (10.7%) | 58 (17.1%) | 45 (13.9%) |
| MZF | 559 (39.4%) | 470 (40.3%) | 339 (39.1%) | 316 (41.3%) | 229 (34.5%) | 98 (36.0%) | 225 (36.9%) | 143 (36.4%) | 129 (37.9%) | 108 (33.3%) |
| DZM | 56 (3.9%) | 41 (3.5%) | 25 (2.9%) | 18 (2.4%) | 37 (5.6%) | 14 (5.1%) | 37 (6.1%) | 21 (5.3%) | 17 (5.0%) | 9 (2.8%) |
| DZF | 293 (20.7%) | 233 (20.0%) | 190 (21.9%) | 167 (21.9%) | 131 (19.7%) | 58 (21.3%) | 113 (18.6%) | 94 (23.9%) | 63 (18.5%) | 68 (21.0%) |
| DZO | 211 (14.9%) | 176 (15.1%) | 128 (14.8%) | 121 (15.8%) | 114 (17.2%) | 43 (15.8%) | 108 (17.7%) | 62 (15.8%) | 50 (14.7%) | 63 (19.4%) |
| Unknown zygosity | 126 (8.9%) | 105 (9.0%) | 79 (9.1%) | 73 (9.6%) | 70 (10.5%) | 22 (8.1%) | 51 (8.4%) | 31 (7.9%) | 23 (6.8%) | 31 (9.6%) |
| Total | 1418 | 1166 | 867 | 764 | 664 | 272 | 609 | 393 | 340 | 324 |

**Table 4.**
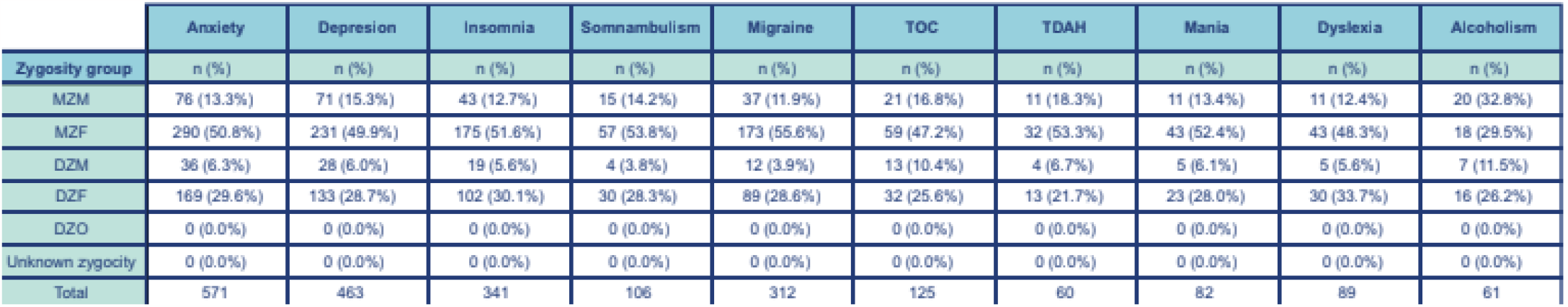
The mental health conditions with the greatest occurrence rates among twins recorded on TwinsMX.

### Advances in the Magnetic Resonance Imaging Study

To date, we have collected Magnetic Resonance data from more than 270 twins, mainly from the metropolitan areas of Queretaro and Mexico City. In **Figure 5** representative images of the high-resolution and high-contrast anatomical images are shown. Functional MRI includes a resting-state acquisition (10 minutes long) and the execution of a visuospatial working memory task, previously designed by our group to assess individual differences in performance as a function of diversity and frequency of leisure activities in Mexican young adults (Ruiz-Contreras et al., 2012). A representative image of the functional activation maps of the working memory task can be seen in the **Figure 6**, active regions include the typical pattern identified in previous work, including the central executive network (*i*.*e*., lateral frontal, parietal cortex, and occipital regions). Finally, a representative analysis of a magnetic resonance spectroscopy of the medial parietal cortex is shown in **Figure 7**. We can see the main peaks associated with NAA; tCr, Cho, Glx and mI, these spectra are obtained by a Point RESolved Spectroscopy (PRESS) sequence. GABA and GSH estimation is performed using the Hadamard Encoding and Reconstruction of MEGA-Edited Spectroscopy sequence (HERMES; Saleh et al., 2016).

**Figure 5.**
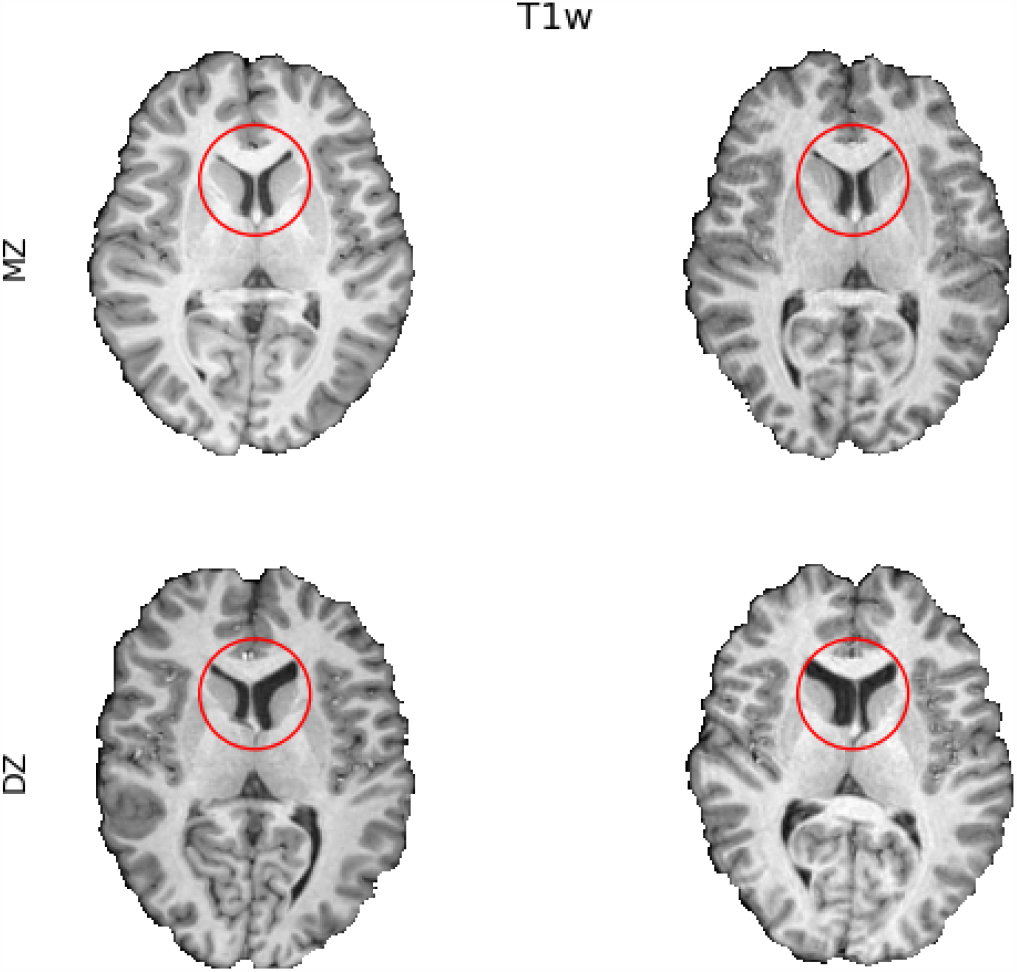
Example high-resolution and high-contrast T1w images. The slices are from a set of MZ and DZ twins. Ventricle shape and volume appear to be more similar between identical twins than fraternal twins.

**Figure 6.**
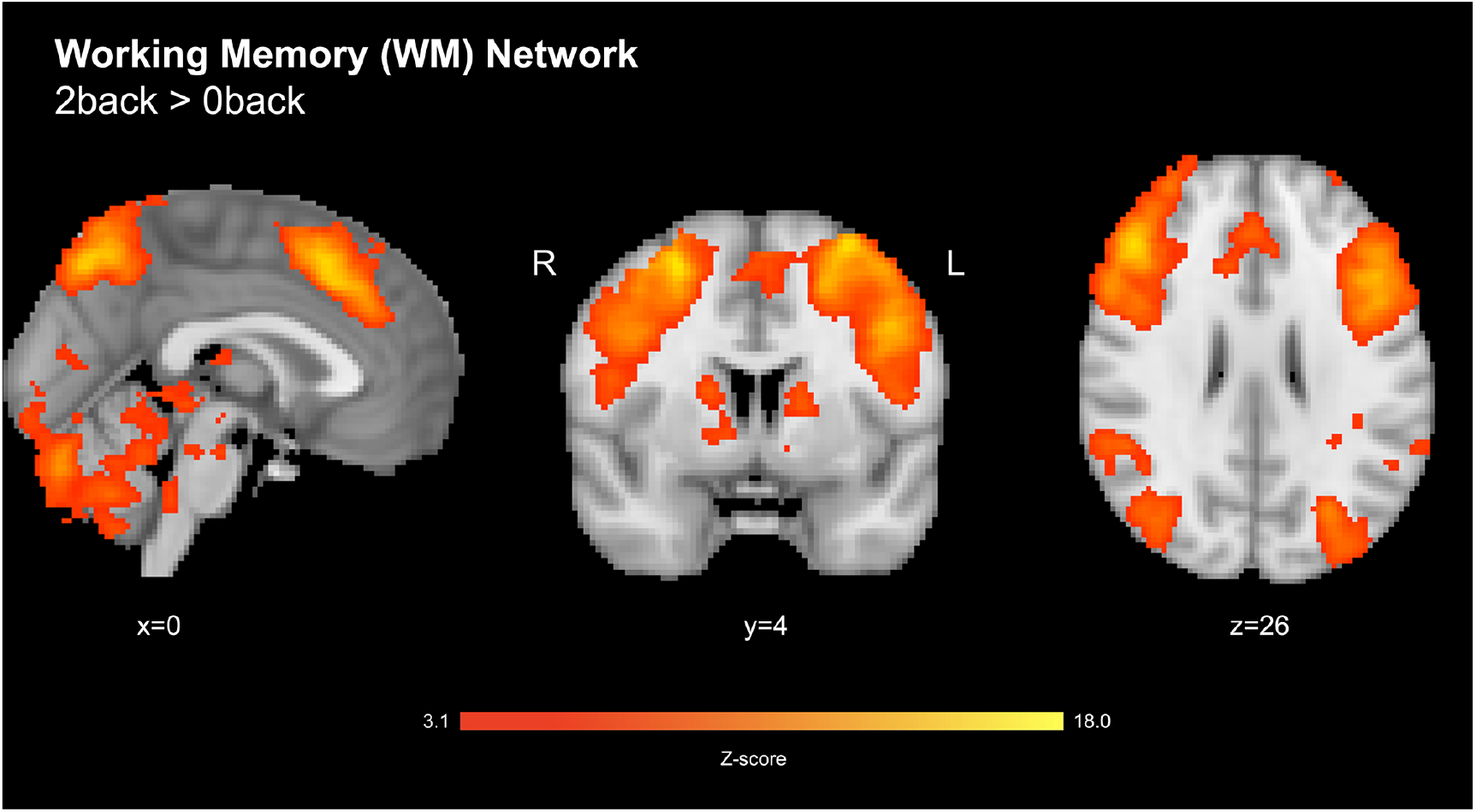
Working memory network. Contrast between 2-back and 0-back tasks. Representative figure (n=28) of the working memory which includes: the precuneus (PCu), the paracingulate gyri (PCG), cerebellum, bilateral medial frontal gyri (FMG), bilateral caudate nuclei (CN), bilateral occipital cortex (OC), dorsolateral prefrontal cortex (DLPFC), and operculum. Image thresholded z-stats. Z>3.1, cluster-corrected p <0.05.

**Figure 7.**
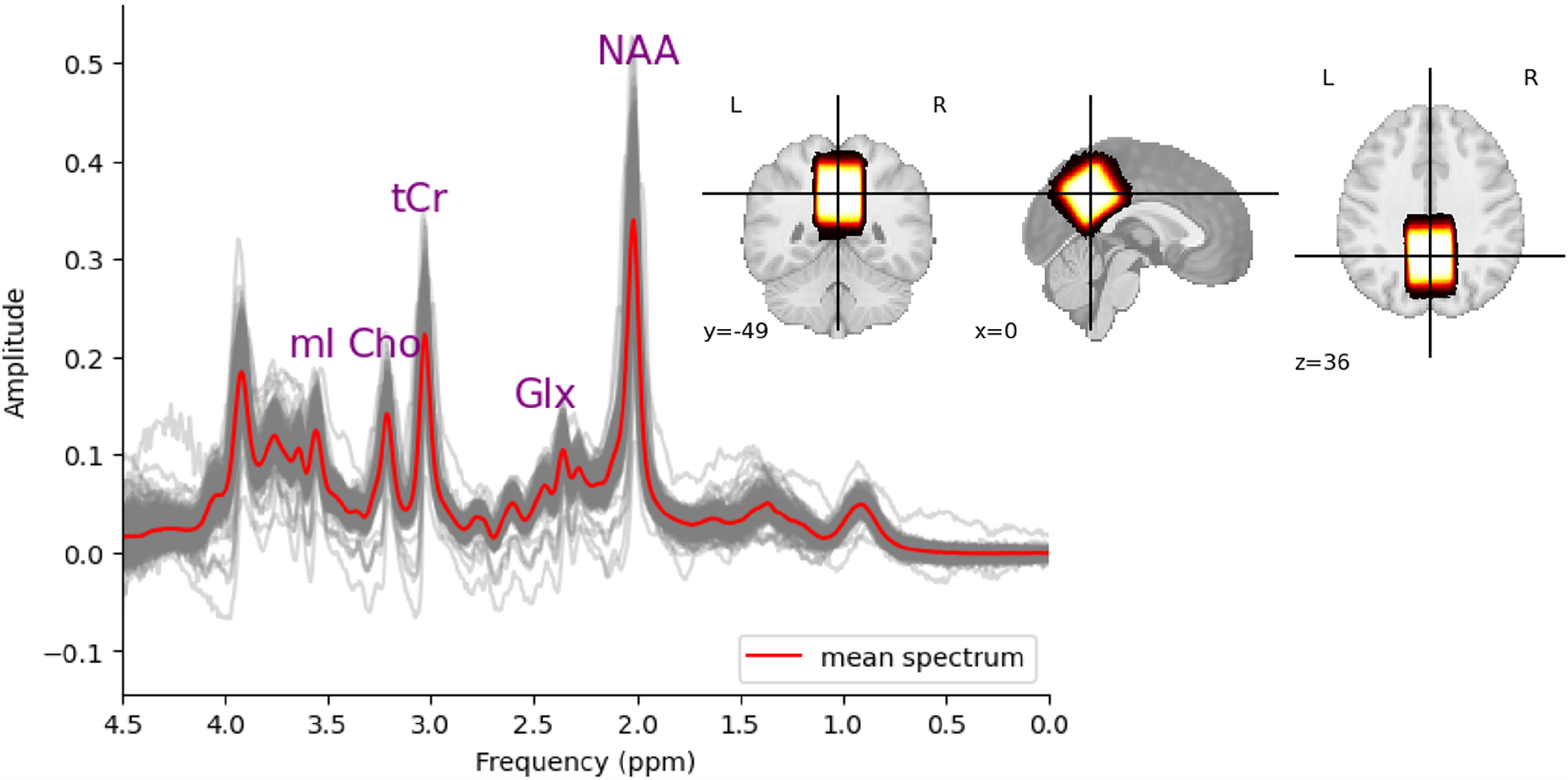
Magnetic resonance spectroscopy data from the medial parietal cortex. In gray the individual spectra of each participant, and in red the average spectrum of the sample. The main peaks of NAA, Glx, tCr, Cho, and mI are labeled. The overlap of subjects’ voxel placements is shown on the right side, where brighter colors represent a higher overlap.

## Discussion

Twin studies have been instrumental in uncovering the underlying causes of a wide range of complex traits and diseases. The insights gained through these studies have also been invaluable in the development of new therapies and treatments. The creation of a twin registry in Mexico, TwinsMX, is particularly important given the unique genetic makeup of the Mexican population, which is a mixture of various ethnic groups. The geographic origin of the registered twins on the TwinsMX platform reflects regional differences in population density. In general, the number of records is in line with the population of each state; however, there are notable exceptions, such as the state of Querétaro. This state is not one of the most heavily populated, yet it has a high number of records. This may be because many campaigns are carried out there, where MRI studies are also performed. This highlights the need to strengthen the dissemination of information in other states’ media.

Another issue that is worth mentioning is that there are more female participants in the project than male participants. While TwinsMX campaigns are not gender-specific, this pattern should be minimized by encouraging participation of more male twins. However, we must note that the proportion of male twins is known to be lower than that for female twins (https://pubmed.ncbi.nlm.nih.gov/1053246/), and females tend to self-select for online surveys more often than males (https://eric.ed.gov/?id=ED501717). Additionally, the majority of participants have a bachelor’s degree or higher education. This population group may have greater ease of access to equipment and time to complete questionnaires. As such, a recruitment campaign targeting educational groups with fewer records may be necessary to facilitate access to the platform. Finally, it is worth noting that there are a good number of minors (<18 years old) who are already registered on the TwinsMX platform. This indicates that the parents of the twins are willing to participate in the project. This could facilitate access to follow-up visits in future studies.

The TwinsMX registry is gathering information about the most prevalent physical and mental conditions in the Mexican population, offering new perspectives on the nation’s health problems. These findings might guide the creation of focused therapies and treatments for certain disorders. For instance, the twin population’s high frequency of diseases like colitis and being overweight draws attention to the necessity for initiatives to deal with illnesses connected to diet and stress factors in the Mexican population. The increasing incidence of mental health conditions, including anxiety and depression, further supports the need for improving access to mental health care. The health problems impacting the twin population in Mexico may be contrasted with those affecting twin populations in other nations, which is also useful for other purposes. This could lead to fresh insights into the genetics of complex characteristics and illnesses as well as novel discoveries. Additionally, by detecting genetic differences that influence how people respond to diseases and drugs, the registry may help find tailored therapies for people with certain genetic profiles.

TwinsMX is anticipated to significantly advance the field of epidemiological genetics in Mexico and raise our knowledge of the genetic and environmental mechanisms underlying ethnic variation in complex traits and diseases, such as behavioral, psychometric, and anthropometric traits, as well as metabolic, cardiovascular, and mental diseases. An additional objective of TwinsMX is the public outreach of the benefits of twin studies, and to disseminate the scientific findings among participants and the general public. For instance, current TwinsMX results for eye health have shown that astigmatism and myopia in Mexican population has an heritability higher than 60% (see preprint Román-López et al., 2023), which shows that genetic factors are contributing more for these traits.

The TwinsMX project has made significant progress, but there are still several issues that need to be resolved before it can reach its full potential. To continue the recruiting efforts and data collection required to create a comprehensive twin registry, we search for long-term support. The fact that some twins do not complete all of the questionnaires, which might restrict the quantity of data that is acquired, is another project constraint. This demonstrates the necessity of continuing efforts to include and inform twins about the significance of the study and the advantages of taking part. Additionally, TwinsMX only has one branch work team at this time, and it is situated in the interior of the nation. Due to this, it is more challenging to locate twins who reside in more rural places, and it may also be a limitation on the variety of the twin population represented in the registry. Therefore, expanding the research group by creating more branches in other parts of the nation would be advantageous. Additionally, the logistics of transporting and storing biological samples is challenging. Addressing these restrictions, the registry would be more efficient in representing a larger sector of the Mexican population.

## Future perspectives

In the future, it is expected to expand the scope of the project and reach more people through promotion on social networks and collaborations with community organizations. To reach a wider population of twins of different ages, TwinsMX plans to collaborate with schools of various educational levels. This will facilitate sending detailed project explanations to twin students, inviting them to participate and subsequently conducting telephone interviews. The advertising campaign aims to encourage active sharing of digital information. Additionally, TwinsMX will participate in radio shows and interviews to bring study results to the public’s attention. Nationwide twin events will also be organized in the coming years to foster a strong network of connections with twins. The possibility of broadening the focus of the project to include not only physical and mental health but also aspects related to financial health and general quality of life is also being considered. Future waves may consider the integration of health monitoring applications and physical activity monitoring devices to provide a more complete and personalized vision of the health of each user. The data gathered for the project may ultimately be used to improve public policy and assist people and communities in making decisions about their health and well-being.

The TwinsMX registry is anticipated to be a valuable resource for research into complicated diseases and areas of healthy aging in the Mexican community. The resource will be enhanced through increasing data collection, database management, biological sample storage, and statistical quality control. TwinsMX’s founding operations are still supported by a grant from the National Council of Humanities, Science and Technology (CONAHCYT). TwinsMX will continue to collect information from twins through the aforementioned methods for years to come, as well as research new communication technologies and promote content. TwinsMX will explore the possibility of establishing collaborations with researchers from other countries, and compare the findings of Mexican twins with those of other populations. This will provide a more global perspective on the genetics of complex traits and diseases and will lead to discoveries.

The age range of the registration will be widened to include volunteer twins from infancy to adulthood. The precise mapping of genetic risk factors linked to a range of disorders would benefit from the molecular genetic study conducted in the Mexican population. The TwinsMX project has a bright future. The twin registry will expand as recruiting attempts are made, giving researchers a wealth of information to investigate the genetic and environmental factors that impact on complex traits and disorders in the Mexican community.

## Data Availability

All data produced in the present study will be available upon request to the authors.

## Acknowledgments

First, we are extremely grateful to the twins and their families for their active and disinterested involvement with the project. We acknowledge the support and resources from the Universidad Nacional Autónoma de México (National Autonomous University of Mexico), in particular to the Laboratorio Nacional de Imagenología por Resonancia Magnética (LANIREM, translated as the National Laboratory for Magnetic Resonance Imaging), the Laboratorio Avanzado de Visualización Científica (LAVIS, Advanced Laboratory for Scientific Visualization), and the Laboratorio de Neurogenómica Cognitiva. We also thank Erick H. Pasaye Alcaraz, Leopoldo González Santos, Luis Alberto Aguilar Bautista, Jair García Sotelo for their support.

## Financial support

TwinsMX is supported by funding from the National Council for Humanities, Science and Technology of Mexico (CONAHCYT, Grant no. 6390). A.M.-R.’s laboratory is supported by funding from the Support Program for Research and Technological Innovation Projects of the National Autonomous University of Mexico (PAPIIT-UNAM; Grant no. IA206517-IA201119); A.E.R.-C.’s laboratory is supported by PAPIIT-UNAM (Grant no. IN217221). S.A. is supported by PAPIIT-UNAM (Grant no. IN208622). M.E.R. thanks the support of the Rebecca L. Cooper Medical Research Foundation through an Al & Val Rosenstrauss Fellowship (F20231230).

## Notes

### Competing Interest Statement

The authors have declared no competing interest.

### Author Declarations

Research Ethics Committee of the Institute of Neurobiology at Universidad Nacional Autónoma de México gave ethical approval for this work.

## References

Agudelo-Botero, M., Giraldo-Rodríguez, L., Rojas-Russell, M., González-Robledo, M. C., Balderas-Miranda, J. T., Castillo-Rangel, D., & Dávila-Cervantes, C. A. (2021). Prevalence, incidence and years of life adjusted for disability due to depressive disorders in Mexico: Results of the Global Burden of Disease Study 2019. Journal of Affective Disorders Reports, 6, 100206.

Alcauter, S., García-Mondragón, L., Gracia-Tabuenca, Z., Moreno, M. B., Ortiz, J. J., & Barrios, F. A. (2017). Resting state functional connectivity of the anterior striatum and prefrontal cortex predicts reading performance in school-age children. Brain and Language, 174, 94–102.10.1016/j.bandl.2017.07.007.

Bodner M., Perego U.A., Gomez J.E., Cerda-Flores R.M., Rambaldi Migliore N., Woodward S.R., Parson W., Achilli A. (2021) The Mitochondrial DNA Landscape of Modern Mexico. Genes (Basel). 2021 Sep 21;12(9):1453.

Busch, A. M., Pan, Y., & Martin, E. (2015). RedCap in the psychology department: a case study of successful adoption and use. Frontiers in Psychology, 6, 1571.

Chen, C., & Liang, K. Y. (2013). Twin registries in Asia. Twin Research and Human Genetics, 16(6), 944–951.

Chen, X., & Liang, K. Y. (1999). Genetic and environmental influences on schizophrenia: a study of monozygotic twins in China. Schizophrenia Research, 39(3), 199–206.

Cortesi, P. A., Fornari, C., Madotto, F., Conti, S., Naghavi, M., Bikbov, B., … Mantovani, L. G. (2021). Trends in cardiovascular diseases burden and vascular risk factors in Italy: The Global Burden of Disease study 1990–2017. European Journal of Preventive Cardiology, 28(4), 385–396.

De Oliveira TC, Secolin R and Lopes-Cendes I (2023) A review of ancestrality and admixture in Latin America and the caribbean focusing on native American and African descendant populations. Front. Genet. 14:1091269, 10.3389/fgene.2023.1091269.

Derom, C., Vlietinck, R., Loos, R., Derom, R., & Thiery, E. (2017). Twin registries in Europe: a review. Twin Research and Human Genetics, 20(6), 511–517.

Dupuis, J., Neale, B. M., Farrall, M., Bowden, D. W., Franks, P. W., Gao, X., … & Wilson, J. F. (2004). A genome-wide scan for hypertension in African Americans: the NHLBI Family Blood Pressure Program. Hypertension, 44(3), 345–351.

Evers, A., & Schouten, E. G. (2018). Reducing data entry errors in a longitudinal study using RedCap: A case study. PloS one, 13(6), e0198205.

Gleichgerrcht, E., Ibáñez, A., Roca, M., & Torralva, T. (2019). Affective Theory of Mind in patients with behavioral variant frontotemporal dementia: A longitudinal study combining experimental and voxel-based morphometry measures. Journal of Neuropsychology, 13(2), 204–224.

Gracia-Tabuenca, Z., Moreno, M. B., Barrios, F. A., & Alcauter, S. (2018). Hemispheric asymmetry and homotopy of resting state functional connectivity correlate with visuospatial abilities in school-age children. NeuroImage, 174, 441–448.10.1016/j.neuroimage.2018.03.051.

Grier, Mark D., Zimmermann, Jan., Heilbronner Sarah R. (2020). Estimating Brain Connectivity With Diffusion-Weighted Magnetic Resonance Imaging: Promise and Peril, Biological Psychiatry: Cognitive Neuroscience and Neuroimaging. Volume 5. Issue 9.

Harvey, L. A. (2018). REDCap: Web-based software for all types of data storage and collection. Spinal Cord, 56, 625.10.1038/s41393-018-0169-9.

Hur, Y.-M., & Craig, J. M. (2013). Twin registries worldwide: An important resource for scientific research. Twin Research and Human Genetics, 16, 1–12.10.1017/thg.2012.147.

Ikeda A, Iso H, Yamagishi K, Inoue M, Tsugane S. (2009). Blood pressure and the risk of stroke, cardiovascular disease, and all-cause mortality among Japanese: the JPHC Study. American Journal of Hypertension 22(3):273–280.

International Organization for Standardization. (2020). ISO 3166-2:2020 Codes for the representation of names of countries and their subdivisions — Part 2: Country subdivision code. https://www.iso.org/iso-3166-country-codes.html

Juárez-Cedillo T, Zuñiga J, Acuña-Alonzo V, Pérez-Hernández N, Rodríguez-Pérez JM, Barquera R, Gallardo GJ, Sánchez-Arenas R, García-Peña Mdel C, Granados J, Vargas-Alarcón G. (2008) Genetic admixture and diversity estimations in the Mexican Mestizo population from Mexico City using 15 STR polymorphic markers. Forensic Sci Int Genet. 2008 Jun;2(3):e37–9.

Leon-Apodaca, A. V., Chiu-Han, E., Ortega-Mora, I., Román-López, T. V., Caballero-Sánchez, U., Aldana-Assad, O., Campos, A. I., Cuellar-Partida, G., Ruiz-Contreras, A. E., Alcauter, S., Rentería, M. E., Medina-Rivera, A. TwinsMX Uncovering the basis of health and disease in the Mexican population. Twin Research and Human Genetics, 2019;22(6):611–616. 74.

Liu S, Li Y, Zeng X, et al. (2019). Burden of Cardiovascular Diseases in China, 1990-2016: Findings From the 2016 Global Burden of Disease Study. JAMA Cardiology 4(4):342–352.

Loehlin, J. C. (2022). “History of Twin Studies” in Tarnoki, A. D. (Ed.), Tarnoki, D. L. (Ed.), Harris, J. R. (Ed.), Segal, N. L. (Ed.), Twin Research for Everyone: From Biology to Health, Epigenetics, and Psychology (pp. 3–6) Academic Press.

Luciano, M., Harris, S. E., Payton, A., & Starr, J. M. (2015). Twin studies in psychiatry: a review. American Journal of Medical Genetics Part B: Neuropsychiatric Genetics, 168B(7), 485–494.

Martin, A. R., Kanai, M., Kamatani, Y., Okada, Y., Neale, B. M., & Daly, M. J. (2019). Clinical use of current polygenic risk scores may exacerbate health disparities. Nature Genetics, 51, 584–591.10.1038/s41588-019-0379-x.

Martinez-Fierro, M., Beuten, J., Leach, R. et al. (2009) Ancestry informative markers and admixture proportions in northeastern Mexico. J Hum Genet 54, 504–509.

Medina-Mora, M., Borges, G., Benjet, C., Lara, C., & Berglund, P. (2007). Psychiatric disorders in Mexico: Lifetime prevalence in a nationally representative sample. The British Journal of Psychiatry, 190(6), 521–528, 10.1192/bjp.bp.106.025841.

Moreno-Estrada, A., Gignoux, C. R., Fernández-López, J. C., Zakharia, F., Sikora, M., Contreras, A. V., Bustamante, C. D. (2014). Human genetics. The genetics of Mexico recapitulates Native American substructure and affects biomedical traits. Science, 344, 1280–1285.

Olsson, T., Barcellos, S., et al. (2013). Understanding participation in genetic research among patients with multiple sclerosis: The influences of ethnicity, gender, education, and age. Multiple Sclerosis Journal, 19(1), 75–82, 10.1177/1352458512456277.

Organización Mundial de la Salud. (2019). Prevalencia de diabetes y factores de riesgo en adultos.

Ordoñana J. R, Rebollo-Mesa I, Carrillo E, Colodro-Conde L, García-Palomo F. J, González-Javier F, Sánchez-Romera J. F, Aznar Oviedo J. M, de Pancorbo M. M, Pérez-Riquelme F. The Murcia Twin Registry: a population-based registry of adult multiples in Spain. Twin Res Hum Genet. 2013 Feb;16(1):302–6. doi: 10.1017/thg.2012.66. Epub 2012 Oct 9. PMID: 23046559.

Peterson, R. E., Kuchenbaecker, K., Walters, R. K., Chen, C.-Y., Popejoy, A. B., Periyasamy, S., … Duncan, L. E. (2019). Genome-wide association studies in ancestrally diverse populations: Opportunities, methods, pitfalls, and recommendations. Cell, 179, 589–603.10.1016/j.cell.2019.08.051.

Petrill, S. A., Plomin, R., & DeFries, J. C. (2013). Twin studies of cognitive abilities: a review. Twin Research and Human Genetics, 16(6), 952–959.

Plomin, R., DeFries, J. C., Knopik, V. S., & Neiderhiser, J. M. (2014). Twin studies in the 21st century: what have we learned and where are we going? Twin Research and Human Genetics, 17(6), 705–715.

Ruiz-Linares, A., Adhikari, K., Acuña-Alonzo, V., Quinto-Sanchez, M., Jaramillo, C., Arias, W., Fuentes, M., … Gonzalez-José, R. (2014). Admixture in Latin America: geographic structure, phenotypic diversity and self-perception of ancestry based on 7,342 individuals. PLoS Genet, 10(9), 10.1371/journal.pgen.1004572.

Ruiz-Contreras, A. E., Soria-Rodríguez, G., Almeida-Rosas, G. A., García-Vaca, P. A., Delgado-Herrera, M., Méndez-Díaz, M., & Prospéro-García, O. (2012). Low diversity and low frequency of participation in leisure activities compromise working memory efficiency in young adults. Acta Psychologica, 139, 91–96.10.1016/j.actpsy.2011.10.011.

Saleh, M. G., Oeltzschner, G., Chan, K. L., Puts, N. A., Mikkelsen, M., Schär, M., … & Edden, R. A. (2016). Simultaneous edited MRS of GABA and glutathione. Neuroimage, 142, 576–582.

Santomauro, D. F., Mantilla Herrera, A. M., Shadid, J., Zheng, P., Ashbaugh, C., Pigott, D. M., Ferrari, A. J. (2021). Global prevalence and burden of depressive and anxiety disorders in 204 countries and territories in 2020 due to the COVID-19 pandemic. The Lancet, 398(10312), 1700–1712.

Secretaría de Salud de México. (2016). Estrategia Nacional de Prevención y Control de Enfermedades No Transmisibles.

Tuppin, P., Rivière, S., Rigault, A., Tala, S., Drouin, J., Pestel, L., … Fagot-Campagna, A. (2016). Prevalence and economic burden of cardiovascular diseases in France in 2013 according to the national health insurance scheme database. Archives of Cardiovascular Diseases, 109(6-7), 399–411.

Verweij, K. J. H., Agrawal, A., & Kendler, K. S. (2018). Twin studies in addiction research: a review. Addiction, 113(4), 677–688.

Wojcik, G. L., Graff, M., Nishimura, K. K., Tao, R., Haessler, J., Gignoux, C. R., … Carlson, C. S. (2019). Genetic analyses of diverse populations improves discovery for complex traits. Nature, 570, 514–518.

Wright, A. (2016). REDCap: A tool for the electronic capture of research data. Journal of Electronic Resources in Medical Libraries, 13, 197–201.10.1080/15424065.2016.1259026.

Yoon-Mi H, Leonie H. B., Juan R. O., Jeanette T., Sara A. H., Catherine T., Eivind Y., Christine D., Axel S., Gonneke W. (2019) Twin Family Registries Worldwide: An Important Resource for Scientific Research. Twin Res Hum Genet. 2019 Dec; 22(6): 427–437. doi: 10.1017/thg.2019.121

